# Does domestic violence legislation reduce permissive attitudes about intimate partner violence? Longitudinal evidence from men and women from 61 countries

**DOI:** 10.1101/2023.10.23.23297413

**Authors:** Robin A Richardson, Alexandria R Hadd, Nuria Rodriguez-Planas, Katjana Wiederkehr, Farheen Jamshed, Tarik Benmarhnia, Cari Jo Clark

## Abstract

**Background:** Intimate partner violence (IPV) is highly prevalent and has substantial implications for women’s health. Changing IPV attitudes is one pathway to reduce IPV. While evidence suggests that interventions targeting individuals may change IPV attitudes, the effect of wider-scale interventions, such as legislation, remain unknown.

**Methods:** We used individual-level IPV attitudes information collected between 1997 and 2020 by the Demographic and Health Surveys (DHS) and the Multiple Indicator Cluster Surveys (MICS), which we linked with national-level domestic violence (DV) legislation information. We evaluated the effect of adoption of DV legislation on changes in IPV attitudes using a difference-in-differences study design that controlled for time-varying country-level confounding and accounted for staggered timing of legislation adoption.

**Findings:** Our sample included 2,184,047 women from 60 countries and 390,877 men from 40 countries. After controlling for country-level confounders, adoption of DV legislation reduced IPV acceptability among women (average treatment effect among treated (ATT) = -0.07, 95% CI: -0.16, 0.06) and men (ATT = -0.11, 95% CI: -0.22, 0.03) although estimates were imprecise and included the null.

**Interpretation:** DV legislation may reduce permissive IPV attitudes, especially among men, although conclusions should be interpreted cautiously due to imprecise estimates.

**Funding:** Eunice Kennedy Shriver National Institute of Child Health and Human Development (5R00HD104896).

**PUTTING RESEARCH INTO CONTEXT:** *Evidence before this study:* Ecological models identify attitudes about intimate partner violence (IPV) as an important driver of IPV. Almost all empirical research has focused on individual-level determinants of permissive IPV attitudes, and the effect of large-scale factors, including domestic violence (DV) legislation, is not well known. We performed a search in Google Scholar, PubMed, SCOPUS, PsycINFO, and Web of Science, without language restrictions, using search terms relevant to DV legislation (e.g., DV law, IPV legislation, etc.) and IPV attitudes (e.g., intimate partner violence beliefs, attitudes on wife beating, perception about acceptability of violence, etc.). The terminal date of this search was May 23, 2023. We also searched reference lists of relevant studies to widen our search. We found two papers that estimated the association between DV legislation and IPV attitudes, both of which investigated multiple individual and contextual risk factors simultaneously using cross-sectional data from a large selection of low-and middle-income countries (i.e., 41 and 49 countries). Both studies found no association between DV legislation and IPV attitudes among women (first study) or among men and women (second study).

*Added value of this study:* The present study is the first longitudinal assessment of DV legislation on changing IPV attitudes. Prior research was cross-sectional and investigated many determinants simultaneously, which may obscure specific relationships (e.g., by over-controlling for factors that are consequences of changes in IPV attitudes, such as women’s household decision-making) and cannot establish temporal ordering (e. g., if countries with less permissive attitudes about IPV enacted legislation vs. if legislation reduced permissive IPV attitudes). Using comprehensive longitudinal data from men and women from 61 countries, coupled with a difference-in-differences study design that accounts for inherent differences across countries, the present study provides the most rigorous evidence to-date of this relationship.

*Implications for all available evidence:* We found that DV legislation resulted in modest reductions in permissive IPV attitudes among women and slightly more pronounced reduction in permissive IPV attitudes among men, although estimates were imprecise. Given that the majority of IPV is perpetrated by men, coupled with the fact that IPV attitudes is a strong predictor of IPV perpetration, our results suggest that DV legislation may be one strategy to reduce IPV. However, estimates were imprecise, and more research is needed to confirm this relationship. Future research could replicate findings in additional countries and could clarify this relationship among men by exploring specific aspects of DV legislation that may be most relevant for changing IPV attitudes (e.g., criminal sanctions) or evaluate if DV legislation is only effective in the presence of other contextual factors (e.g., equitable gender norms, government institutions with high levels of democracy).

## INTRODUCTION

Intimate partner violence (IPV), highly prevalent in all regions of the world,^1^ is widely acknowledged as a major threat to women’s health and well-being. The ecological model of IPV posits that causes operate on many different levels, ranging from the individual to societal,^2^ and development schemes have accordingly proposed prevention efforts that confront factors on multiple levels.^2, 3^ For example, a newly proposed framework to prevent violence against women put forth by the United Nations and the World Health Organization (RESPECT) suggests a suite of comprehensive interventions targeting individuals (e.g., changing attitudes about violence against women), relationships (e.g., improving conflict management), communities (e.g., creating safe work environments), and structures (e.g., availability of legal and social services for survivors of violence).^3^

Despite the conceptual and development focus on confronting causes of IPV on many levels, empirical research has primarily focused on evaluating factors experienced at the individual or relational level, with few studies focused on larger-scale factors. A 2018 systematic review of longitudinal studies investigating determinants of IPV found 60 studies that evaluated 71 risk and protective factors, but only 7 of these factors were at the community level and none were at the structural level.^4^ Lack of evidence on these broader levels is a critical research gap that limits evidence-informed decision-making about which structural interventions may work, and which may not, to prevent IPV.

Domestic violence (DV) legislation, adopted in over 100 countries since the year 2000,^5^ may transform ideas about IPV attitudes by signaling the unacceptability of violence on a societal level. IPV attitudes are sensitive to various community and structural interventions,^6–10^ offering promising evidence that DV legislation may also affect IPV attitudes. Such changes in IPV attitudes may have substantial implications for IPV: permissive attitudes about IPV is a major risk factor for both IPV perpetration^11–16^ and victimization^11, 16–22^ and has been identified as a risk factor for IPV in the ecological model.^2^ However, to-date only two exploratory studies have investigated the effect of DV legislation on IPV attitudes. These two studies used population-based data from low-and middle-income countries to estimate the cross-sectional association between multiple individual and contextual risk factors simultaneously, finding no overall association between DV legislation and IPV attitudes among men^23^ or women.^23, 24^ However, the lack of observed associations in these studies could be a result of investigating many determinants simultaneously, which may obscure relationships (e.g., by over-controlling for factors that are consequences of changes in IPV attitudes, such as women’s household decision-making), and cannot establish temporal ordering (e. g., if countries with less permissive attitudes about IPV enacted legislation vs. if legislation reduced permissive IPV attitudes). Longitudinal evidence isolating the effect of distinct determinants would provide stronger evidence for (or against) this relationship.

In this study, we help clarify this relationship. Using national-level policy data from the World Bank, linked with individual-level information from the Demographic and Health Surveys and the Multiple Indicator Cluster Surveys, we evaluate the longitudinal association between adoption of DV legislation and changes in attitudes about the acceptability of IPV among men and women. This work uses data over a 23-year period from 2.6 million adults from 61 countries and assesses the differential impact of the policy among specific demographic groups, providing a comprehensive assessment of the impact of DV legislation.

## METHODS

### Study Population

Our study used individual-level IPV attitudes data from the Demographic and Health Surveys (DHS) and the Multiple Indicator Cluster Surveys (MICS). The DHS is a repeated cross-sectional survey, typically administered every 5 years, that collects information from reproductive age women and men in over 100 low-and middle-income countries. Similarly, the MICS is a repeated cross-sectional survey that has been administered in over 100 countries since the mid-1990s to reproductive age women and men. The MICS and DHS survey designers collaborate closely to produce comparable, harmonized information, which can be combined to produce global datasets.^25^ Standardized questions about IPV attitudes were first introduced in the DHS around 1997 and in the MICS around 2005, although administration of the DHS and the MICS, including the selection of specific questions and study participants, was at each countries’ discretion. Therefore, IPV attitudes information was not collected each survey round in all countries, and additionally some countries chose to collect information from only men or women, whereas other countries collected information from both. Therefore, we restricted our sample to countries that collected information about IPV attitudes from men or women in at least two survey rounds and had not adopted DV legislation before the study period (i.e., before 1997). Our final sample included repeated cross-sectional information from 60 countries for the women’s sample and from 40 countries for the men’s sample. We further restricted our sample to participants who were currently or formerly married because some countries did not administer surveys to individuals who had never been married. Both the DHS and the MICS are population-based surveys that select men and women through a multistage sampling procedure, and we applied the weights supplied by the MICS and DHS to derive a longitudinal panel of nationally representative estimates of currently or formerly married men and women.

### Study Measures

#### Domestic violence (DV) legislation

Yearly country-level DV legislation information came from the World Bank’s Women, Business and the Law (WBL) database.^5^ This database was created by WBL staff by surveying professionals with knowledge of local laws (e.g., judges, lawyers, academics, and non-governmental organizations providing gender-related services) about the presence of DV legislation and any amendments to existing law. WBL staff reviewed texts of relevant laws and regulations in each country to verify existence of two aspects of DV legislation: 1) protective orders for victims of domestic violence, and 2) criminal sanctions against domestic violence perpetrators, and they then classified countries fulfilling either criteria as having DV legislation and countries that offered neither as not having legislation.^5^ Major strengths of this database include confirmation of presence of legislation through review of source documents, and the application of the same criteria across time and countries. To our knowledge, this is the only DV legislation database that provides consistent information across time and country.

#### Attitudes about intimate partner violence (IPV)

Attitudes about the acceptability of IPV were captured in the DHS and the MICS by asking men and women if they believe a husband is justified beating his wife in five specific scenarios: 1) if a wife goes out without telling her husband, 2) if a wife neglects the children, 3) if a wife argues with her husband, 4) if a wife refuses to have sex with her husband, or 5) if a wife burns the food. For each question, response options include “agree”, “disagree”, or “don’t know”. We classified a person as having permissive attitudes about IPV if they selected “agree” to at least one of the five scenarios. We aggregated responses to the year and country level using sampling weights provided by the DHS and the MICS, thus providing prevalence estimates representative at the country-level.

#### County-level control variables

Country-level information that may predict adoption of DV legislation and may also affect IPV attitudes were considered potential confounders. Variables include: female labor force participation rates (% of women aged 15 or older in the labor force, provided by the International Labour Organization^26^), literacy rates (% of people aged 15 or older who can read and write, provided by the World Bank^27^), educational attainment gap between men and women (% of men with at least a primary education minus % of women with at least a primary education, calculated from data provided by the World Bank^27^), and gross domestic product (log) (per capita values for gross domestic product expressed in current international dollars converted by purchasing power parity conversion factor and expressed on the natural logarithm scale, provided by the World Bank^27^). Within countries, data was missing for some years. In these instances, we used linear interpolation to estimate missing values in models stratified by country.

### Analytic Approach

We evaluated the effect of DV legislation on men and women’s attitudes about IPV using a difference-in-differences (DID) approach, a common method to estimate the causal effect of a policy change on health. This approach follows units (e.g., countries) over time, some that experienced a policy change (“treatment group”) and some that never adopted a policy (“control group”). Many countries recently adopted DV legislation,^5^ offering a unique opportunity to use this strong study design to evaluate policy effects.

We used a specialized DID estimation procedure, called the fixed effects counterfactual estimator,^28^ that is designed for repeated cross-sectional data and staggered adoption of policy adoption. The fixed effects counterfactual estimator uses an estimation procedure that compares each treated country with its predicted untreated counterfactual, takes the difference between the two to estimate the treatment effect, and then averages these estimated differences to derive the average treatment effect on the treated (ATT).

### Statistical Analysis

We performed analyses with a panel data setup that modeled changes within countries over time. Models controlled for country fixed effects, included a binary variable indicating presence or absence of national-level DV legislation (exposure), and controlled for time-varying country-level confounders, including female labor force participation rate, literacy rate, educational attainment gap between men and women, and gross domestic product (log). Because the estimation procedure used in this analysis (fixed effects counterfactual estimator) uses time since treatment adoption to match treated units with their estimated counterfactual, time since treatment adoption was also accounted for. Due to the relatively small number of treatment units, variances were estimated using a bootstrap procedure.

Contextual factors affect men and women’s IPV attitudes differently,^23^ and therefore we estimated separate effects in sex-stratified models. In addition, we hypothesized that certain demographic groups may be more affected by DV legislation, including younger adults (because they may be more impressionable or more flexible/ready to change their minds), those with higher educational attainment (because they may be more likely to have knowledge of legislation and may have the means to leverage the new law, if needed), and those living in urban areas (because legislation may be more robustly enacted in this setting). Therefore, in additional analyses we estimated effects among specific demographic groups within sex stratum, including by age (15 – 29 years, 30-40 years, and 41 years or older), educational attainment (primary education or less, secondary education or higher), and location (urban, rural). We formally tested heterogeneity among demographic groups with Cochran’s Q test.^29^

## RESULTS

Our study included 2,184,047 women from 60 countries and 390,877 men from 40 countries. Table 1 shows the sample size in each country and the percentage of men and women who believed IPV is justified. Prevalence of the acceptability of IPV was high in our sample among both men (28.0%) and women (43.7%), although prevalence ranged considerably across countries and years. Among men, acceptability ranged from 5.0% in Malawi (in 2014) to 85.1% in Timor-Leste (in 2009), and among women, from 2.1% in Serbia (in 2019) to 94.0% in Afghanistan (in 2011).

**Table 1.**
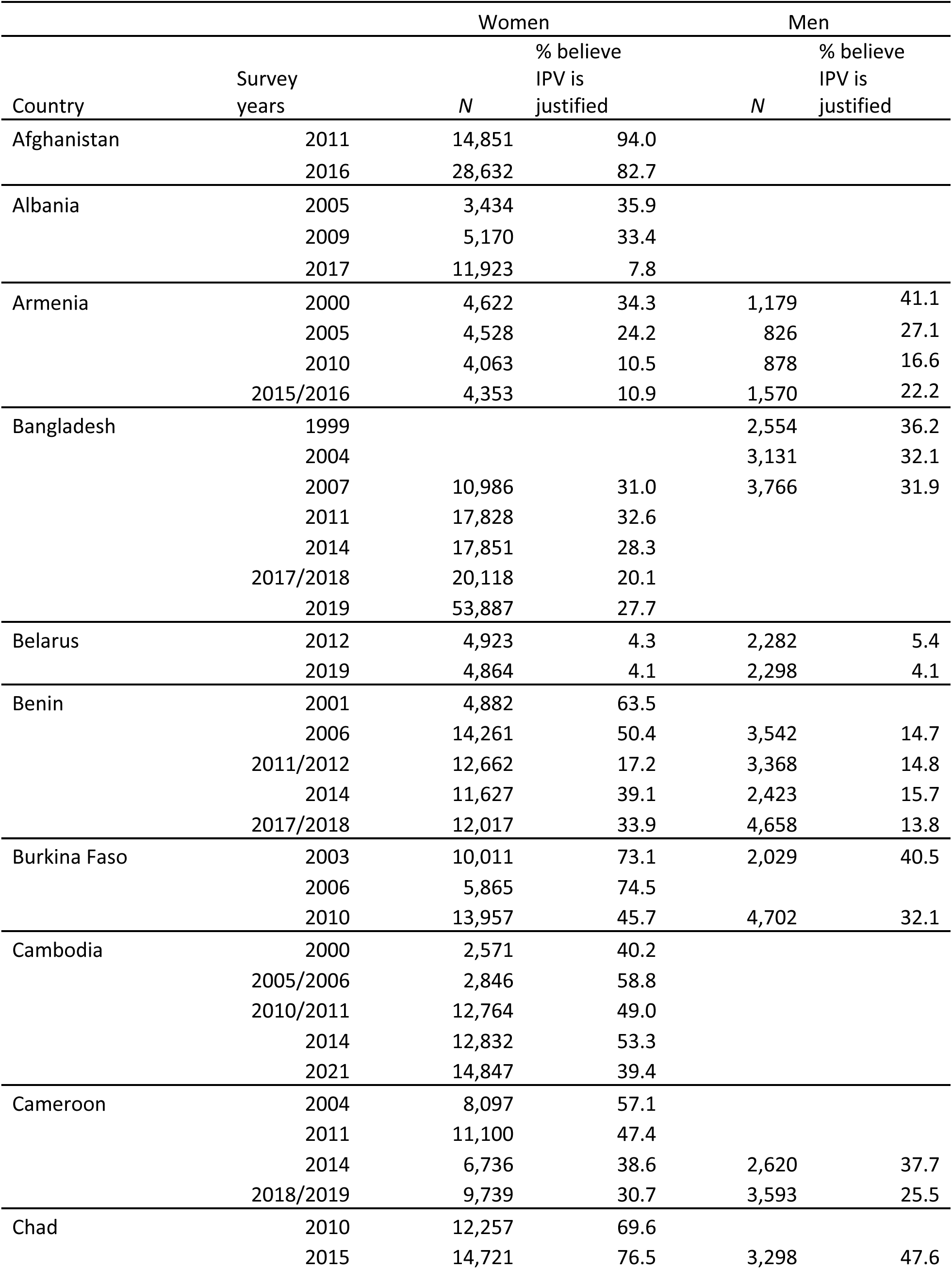
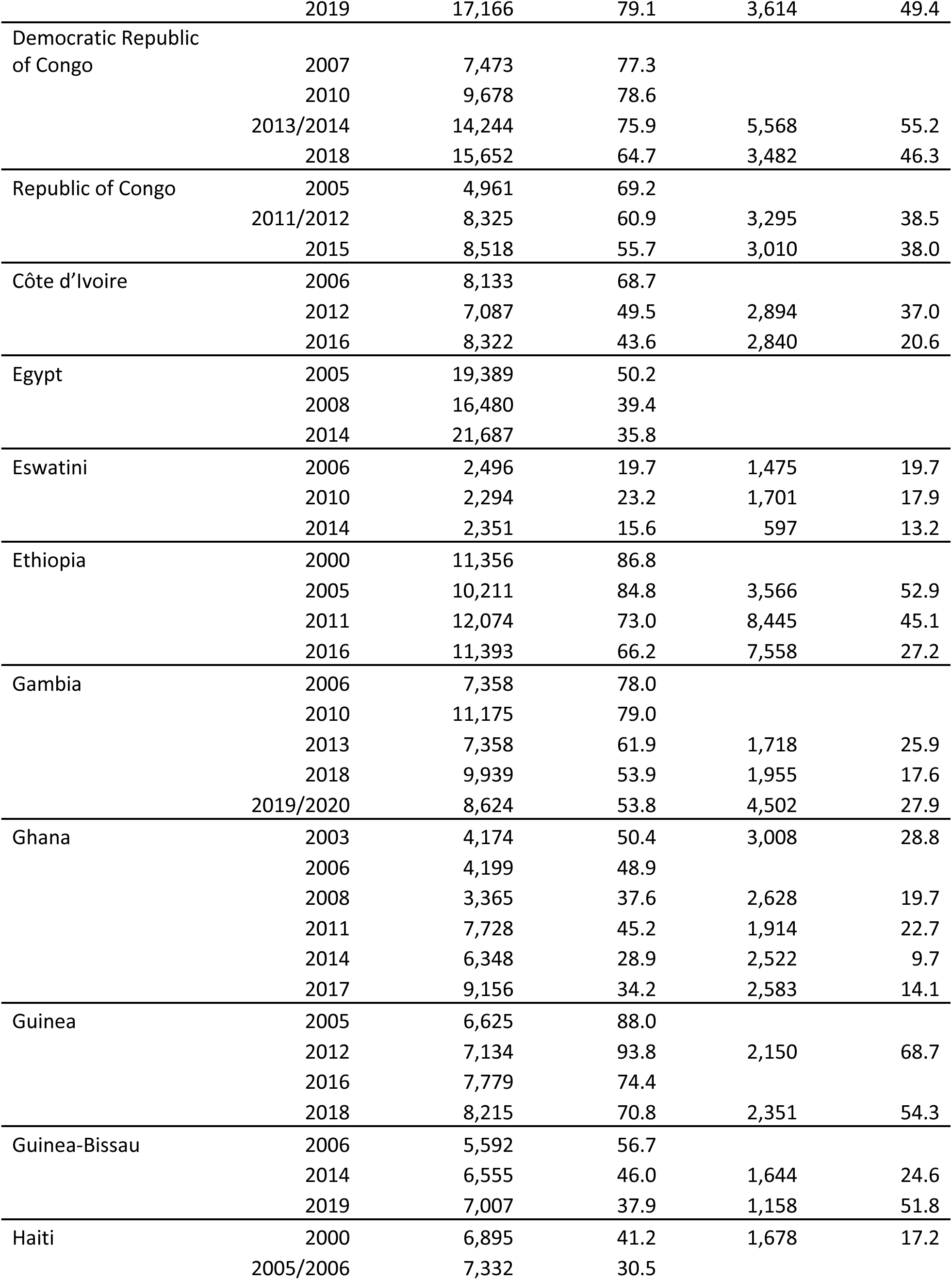
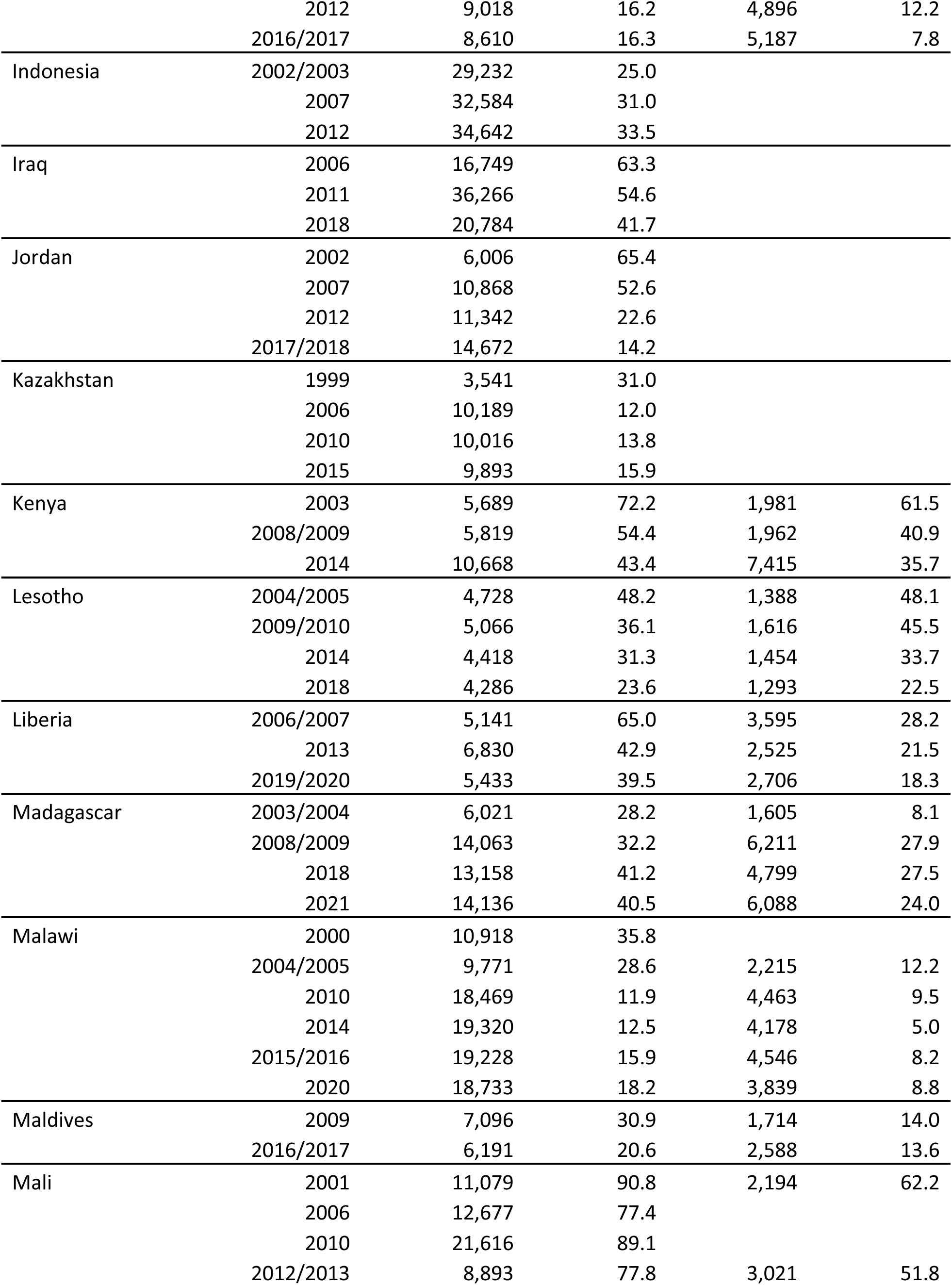
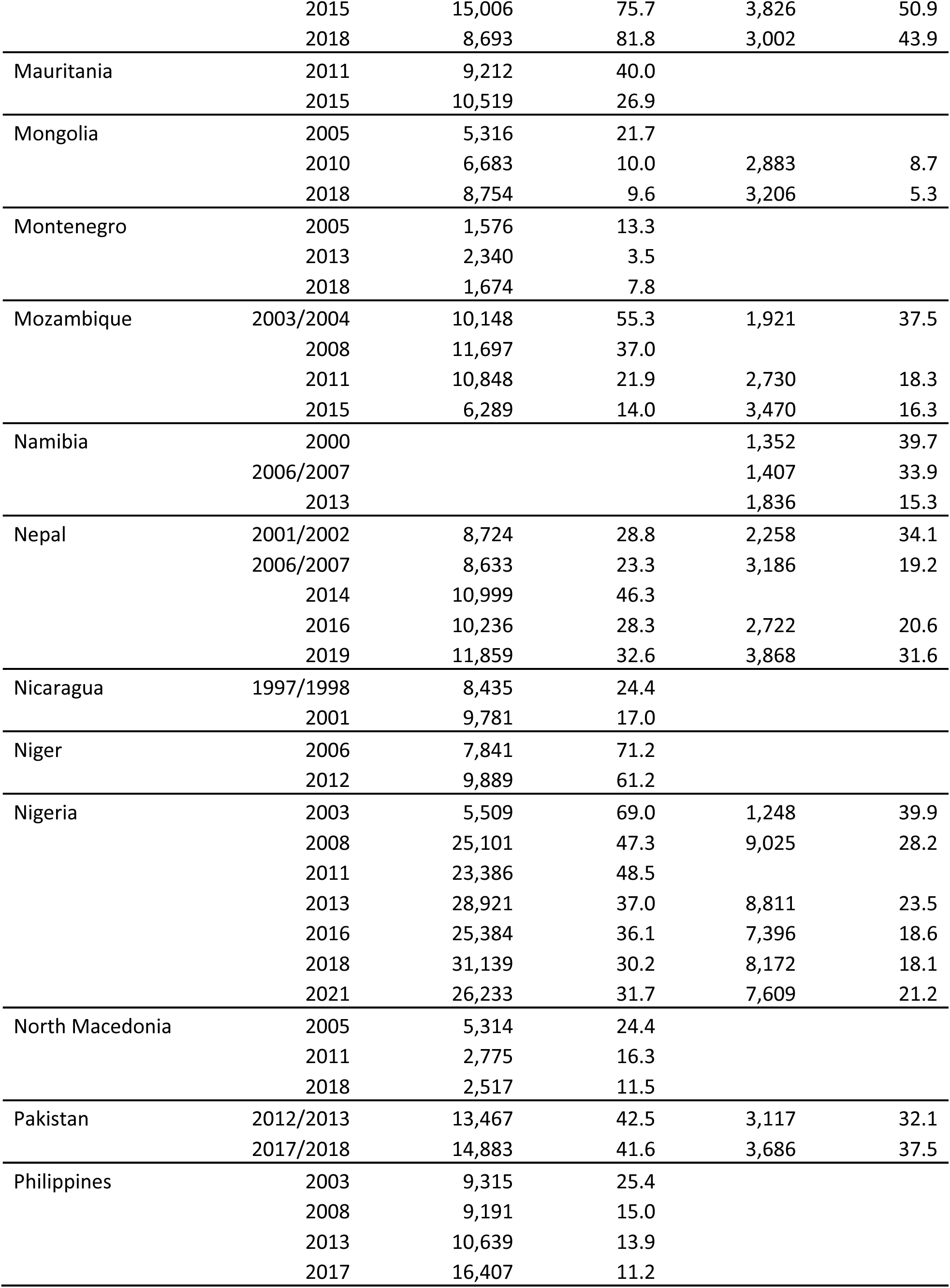
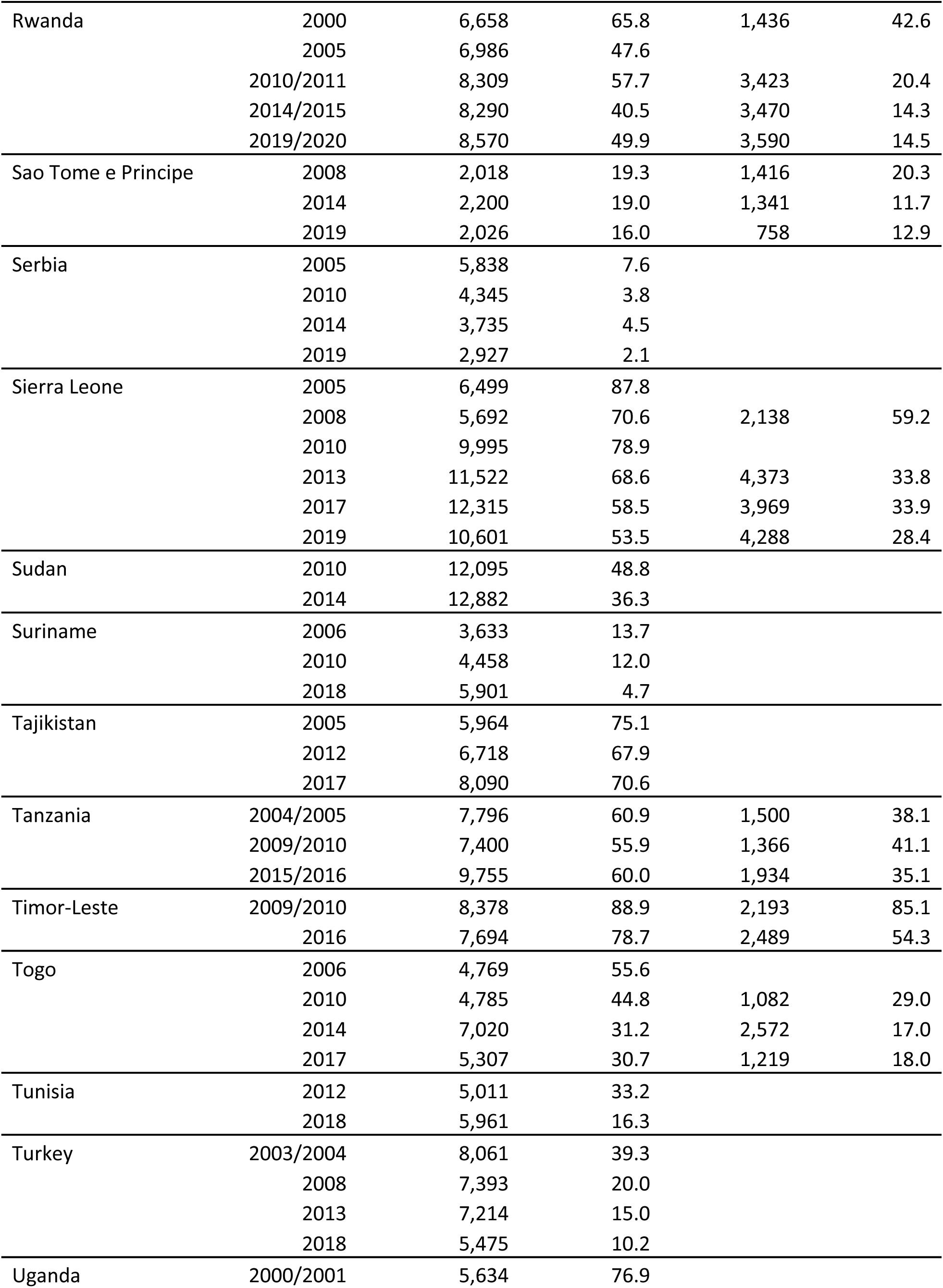
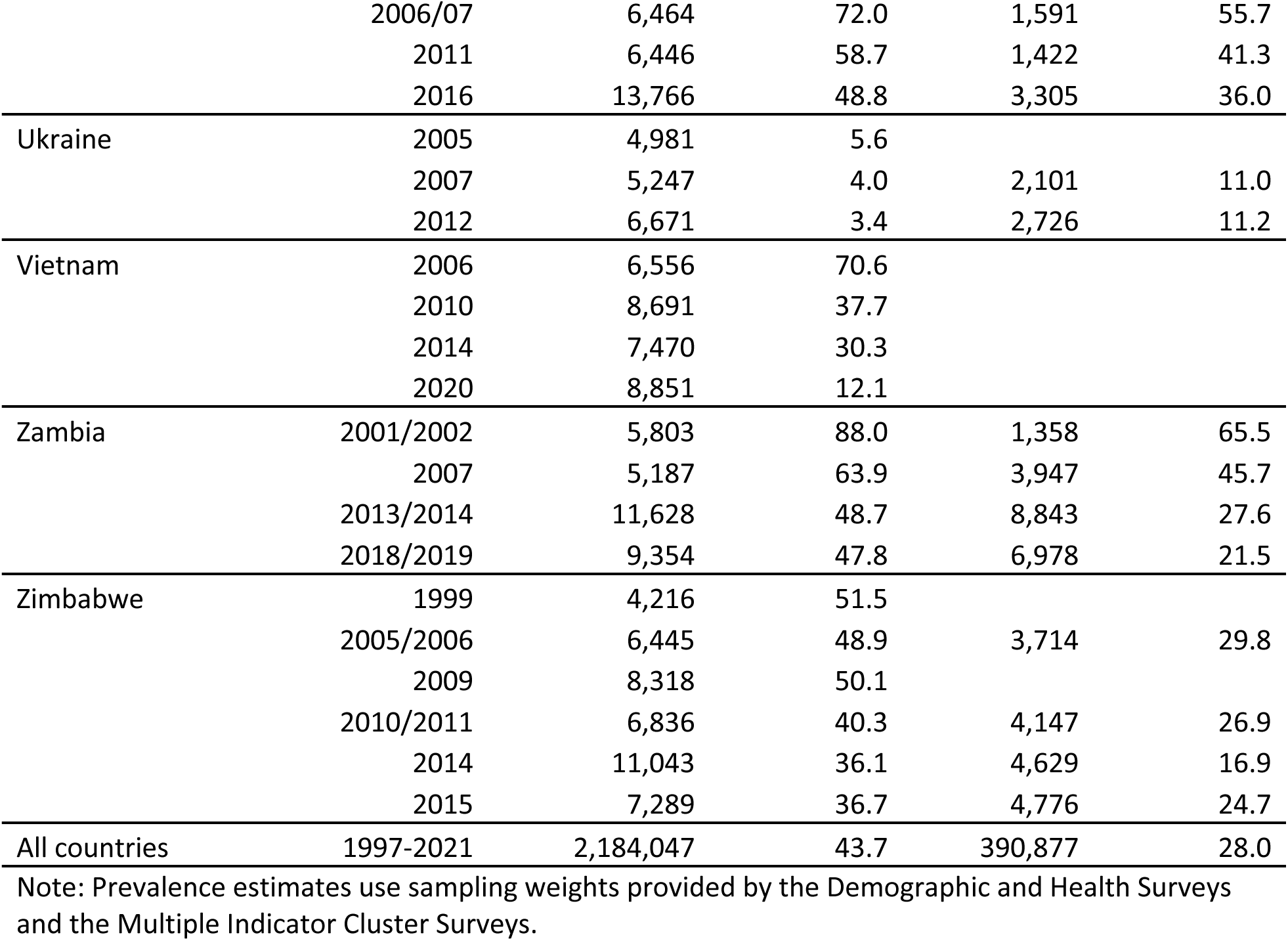
Sample characteristics by country and year.

Many countries adopted DV legislation over the study period. Among the women’s sample, 39 of 60 countries adopted legislation (Figure 1), and among the men’s sample, 25 of 40 countries adopted legislation (Figure 2). There was a strong correlation between presence of DV legislation and lower prevalence of permissive IPV attitudes among both men (*r* = -.28) and women (*r* = -.23).

**Figure 1.**
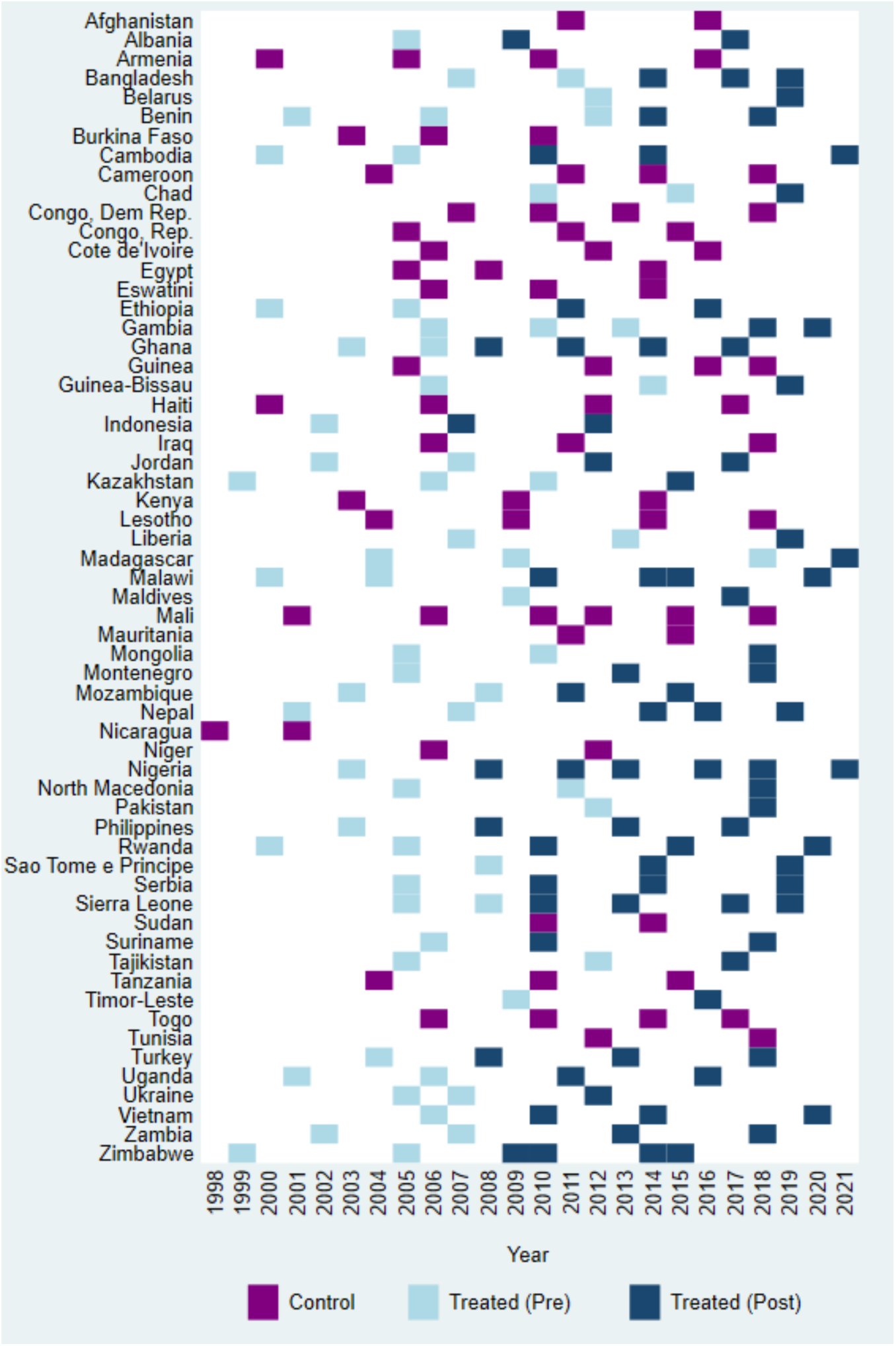
**Domestic violence legislation status and survey availability by country and year for women’s sample.**

**Figure 2.**
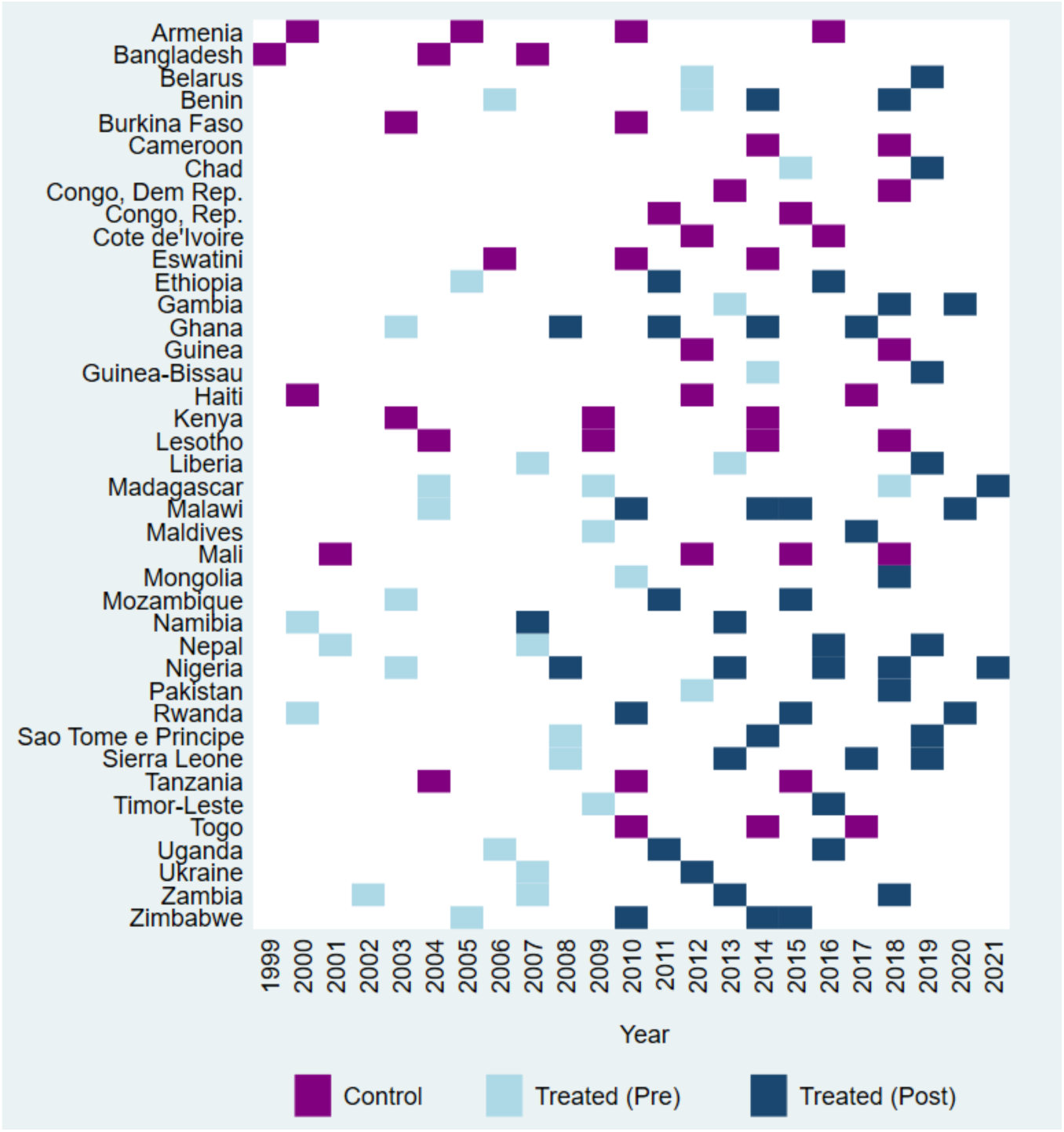
**Domestic violence legislation status and survey availability by country and year for men’s sample**

Figures 3 and 4 show results of the longitudinal analysis evaluating the effect of adoption of DV legislation on changes in IPV attitudes using a DID study design. We found that DV legislation resulted in a 7% reduction in the prevalence of permissive IPV attitudes among women (ATT = -0.07, 95% CI: -0.16, 0.06) and an 11% reduction among men (ATT = -0.11, 95% CI: -0.22, 0.03), after controlling for country-level confounders. However, estimates were imprecise and included the null value. Unadjusted models showed stronger and more precise effects among women (18% reduction in prevalence) (Figure 3), with adjustment for GDP (log) producing the greatest attenuation on women’s effect size. Among men, unadjusted and adjusted estimates had similar effect sizes, although adjusted estimates were more imprecise (Figure 4).

**Figure 3.**
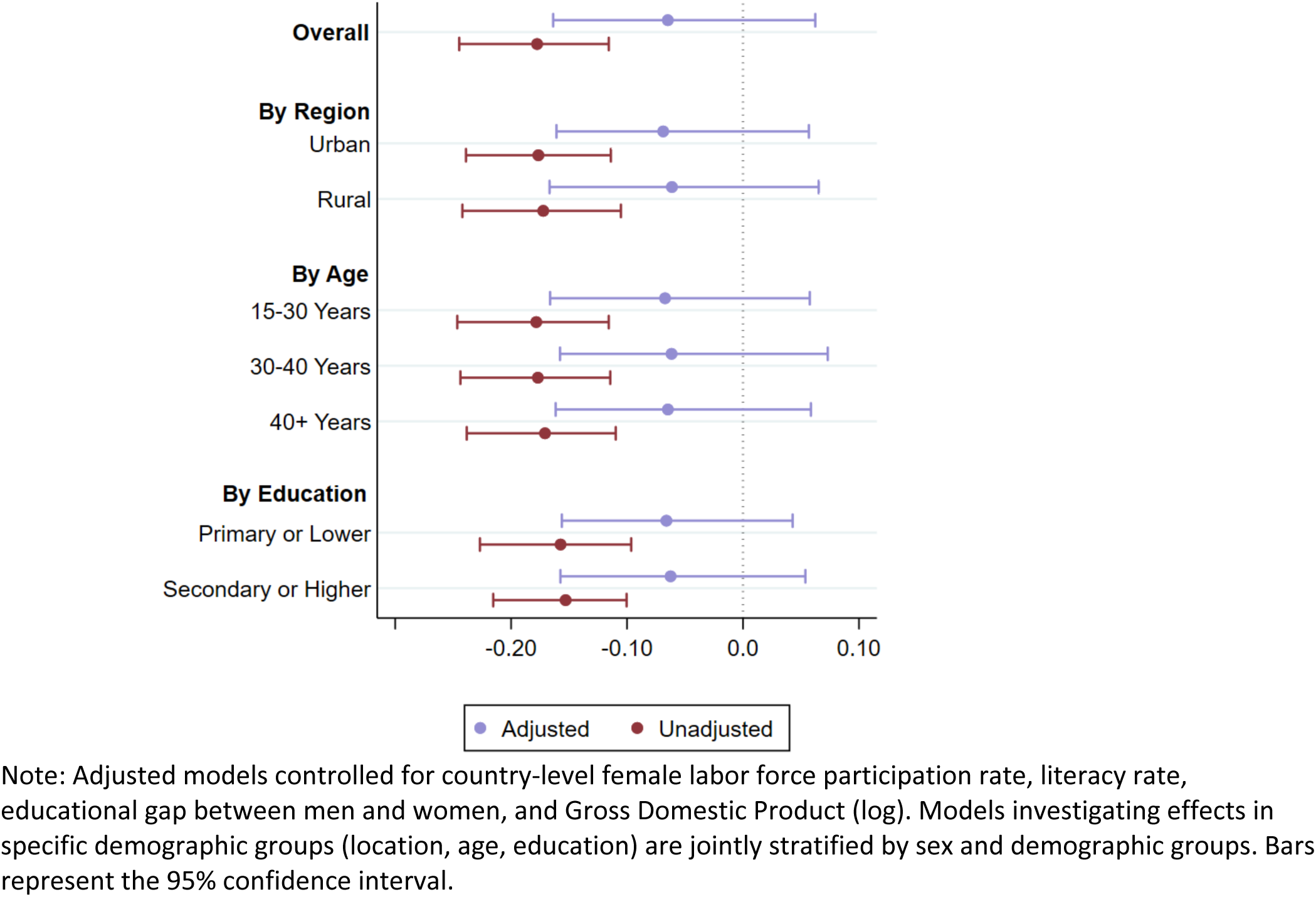
**Effect of adoption of domestic violence legislation on changes in attitudes about IPV among women**

**Figure 4.**
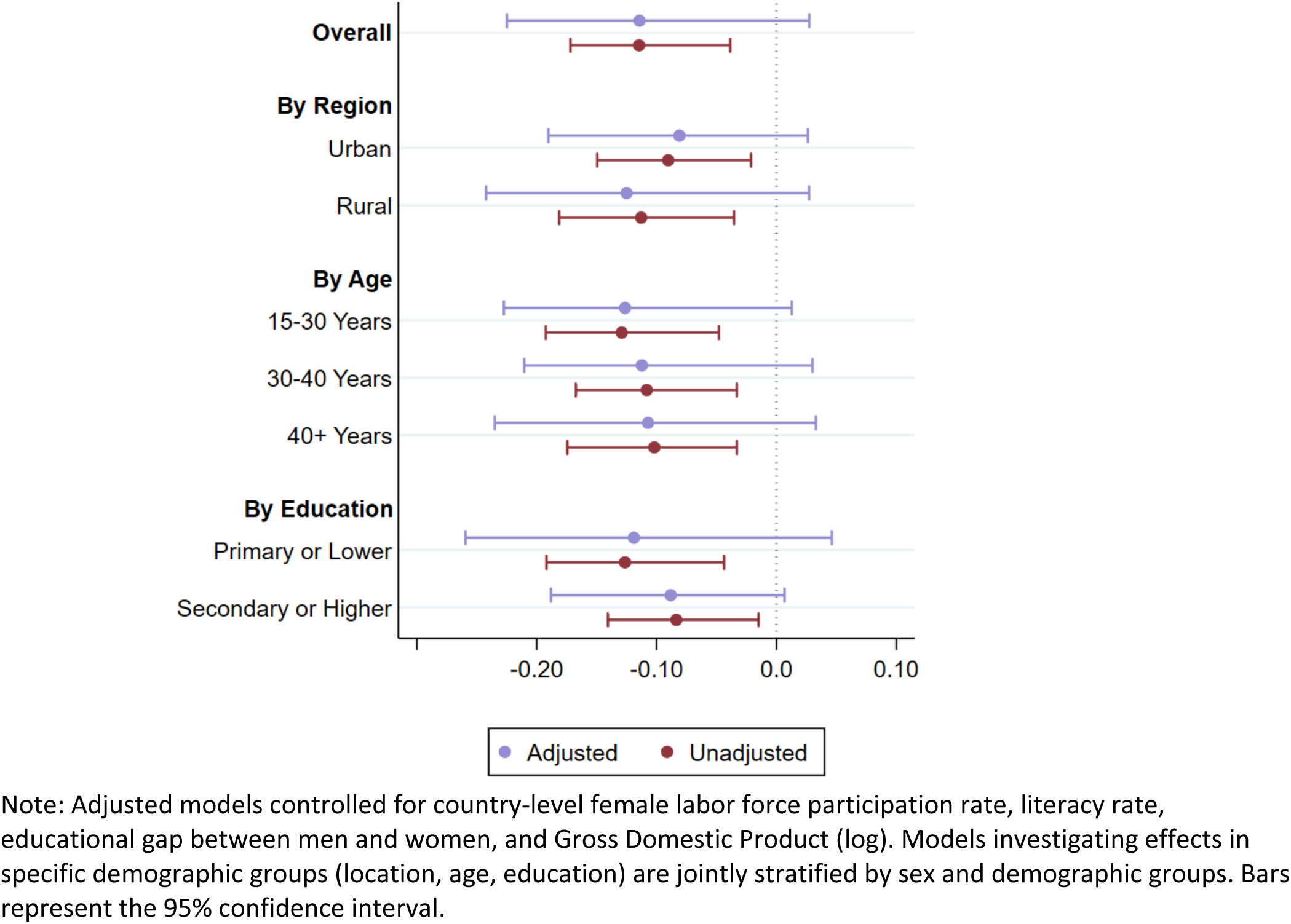
**Effect of adoption of domestic violence legislation on changes in attitudes about IPV among men**

In additional analyses stratified by specific demographic groups hypothesized to be more affected by DV legislation (i.e., urban, younger, or more educated adults) (Figures 3 & 4), we found that effects did not differ among demographic groups in either the men or women’s sample, as indicated by Cochran’s Q test (p >0.5 for all tests within demographic groups).

## DISCUSSION

Development schemes to reduce IPV have strongly emphasized confronting structural determinants, yet empirical evidence on which factors prevent IPV are lacking.^4^ This study helps fill this crucial evidence gap by leveraging longitudinal country-level information, coupled with an innovative, rigorous estimation strategy, to evaluate the effect of one structural factor identified as a major determinate of IPV: men and women’s IPV attitudes. This work provides the most comprehensive assessment to-date of the relationship between DV legislation and permissive IPV attitudes. We found that adoption of DV legislation corresponded to a 7% reduction in the prevalence of permissive IPV attitudes among women and an 11% reduction in prevalence among men. While these results suggest legislation may lead to meaningful reductions in IPV attitudes, estimates were imprecise and confidence intervals included the null, precluding firm conclusions.

The imprecision of estimates, despite a fairly large sample size, may suggest substantial effect heterogeneity due to various contextual factors (e.g., IPV norms, status of women) that may amplify or diminish the effect of DV legislation. The ‘sticky norms problem’ posits that when legislation deviates substantially from norms, judges, police officers, juries, and community members will be less likely to enforce, implement, or support the law, resulting in less effective legislation.^30^ For example, in the US, mandatory arrest laws for perpetrators of DV were not universally adopted due to the belief among some police departments that DV was a relationship issue and not criminal matter,^31^ and extreme laws addressing other social issues, including polygamy and inheritance rights for women, were found to be less effective than more moderate laws that align more closely with local norms.^32^ Accordingly, DV legislation may only be effective in environments where legislation aligns with local norms about violence against women. In addition, the presence of multiple structural factors may be necessary to substantially change IPV attitudes, such as structural determinants that may act upon IPV attitudes more broadly by changing perceptions about women’s role in society (e.g., policies to promote women’s economic empowerment) or promote women’s freedom from violence (e.g., perceptions about the acceptability of divorce). While our study design, a modified difference-in-differences approach, allowed us to rigorously isolate the effect of DV legislation, this approach has limited ability to estimate the joint effect of multiple contextual or structural factors because estimating joint effects requires grouping countries by these additional factors (e.g., the effect of adoption of DV legislation among countries with high permissive IPV attitudes). Given the relatively small number of countries in this study, stratified analyses investigating additional country characteristics was not feasible. Future studies that include more countries, or estimate effects within geographic regions within individual countries, could clarify the influence of contextual environments on DV legislation.

Our study has limitations. First, our study used presence or absence of DV legislation, and this broad measure did not capture granular information about the degree of implementation or the comprehensiveness of enacted legislation. Integrating this information into our evaluation could clarify if DV legislation is effective in specific scenarios. We are not aware of longitudinal databases that systematically collect this information across time and countries, and the creation of new datasets that collect aspects of DV legislation (e.g., criminal sanctions, civil penalties) longitudinally would open up new opportunities to rigorously evaluate the effect of specific aspects of DV legislation. Second, changes in IPV attitudes may occur slowly over time, and the relatively short period of follow-up after adoption of legislation in our study (on average, the women’s and men’s sample included 5.5 years of follow-up) may not allow enough time to detect changes. Thus, future research with more years of follow-up could confirm the relationships observed in this study. Third, the psychometric properties of the IPV attitudes questions used in this study may vary across time or country. These differences may be a potential source of study bias, and a psychometric assessment of these measures could be an additional area of future research. Fourth, our study used data from a selected group of low-and middle-income countries that had information from at least two DHS or MICS surveys, and among these selected countries, our estimation strategy did not use information from countries that always had DV legislation. Thus, the study population was composed of a selected group of countries, and results may not be generalizable to countries beyond this subset.

Despite these limitations, our work provides the most comprehensive assessment to-date on the link between DV legislation and women and men’s IPV attitudes. Understanding specific pathways that may result in reductions in violence – at the individual, relational, community, and structural level – is imperative to effectively reduce violence. Our work helps fill this evidence gap by clarifying the role of one structural determinant, DV legislation, on changing IPV attitudes. Future research could investigate structural determinants that act more broadly upon gender equitable attitudes, which may reveal new pathways for reducing permissive IPV attitudes.

## Contributors

RAR conceptualized the study and wrote the first draft of the report.

RAR, NRP, and TB contributed to the study design.

ARH, KW, and RAR constructed harmonized datasets.

ARH and RAR conducted statistical analysis.

ARH, RAR, CJC, and NRP contributed to the interpretation of results.

FJ and RAR conducted the literature review.

FJ and ARH contributed to the writing of the report.

All authors provided critical feedback of manuscript drafts, and reviewed and approved the final manuscript.

## Data Availability

All individual-level data used in this study is publicly available (with application) through the Demographic and Health Surveys and the Multiple Indicator Cluster Surveys. Country-level policy information is publicly available at the World Bank's website.

## Acknowledgments

This work was supported by the Eunice Kennedy Shriver National Institute of Child Health and Human Development (5R00HD104896). We thank Lori Heise for providing comments on an earlier paper draft.

